# Acute Medical Unit Healthcare workers’ interaction in a developing country: Social Network Analysis

**DOI:** 10.1101/2021.02.05.21250636

**Authors:** Jihane Belayachi, Sarah Benammi, Rhita Nechba Bennis, Naoufel Madani, Redouane Abouqal

## Abstract

**INTRODUCTION:** Social network analysis is used to increase the awareness of leaders about the power of networks, to further catalyze relationships and connections, and to strengthen the capacity of the network to act collectively. We focus on understanding and measuring the communication patterns in an acute medical unit team. We sought to use Social Network Analysis to describe the patterns of communications in teamwork of an acute medical unit.

**METHODS:** Network Analysis was conducted to examine network structure in 58 teamwork professional communication in an AMU. Team members reported the frequency (0 to 10+ times) of professional discussion with every other coworker during the last 48-hoursk, density, degree and betweenness centralization, degree and betweenness centrality; and homophily were calculated. P-value was obtained based on 1000 quadratic assignment procedure QAP permutations of the network. The network analysis was used to construct network maps using multidimentional scaling and generates a visual representation of networks through network diagrams.

**RESULTS:** there were 460 connections (density=28%). The whole network has a moderate degree centralization (37%) and lower betweenness centralization (8%). Three senior physicians, the head nurse, the physiotherapist, the medical secretary and the archive manager were most central in network. There was evidence regarding heterophily in a network indicated by high level of E-I index value 0.34 (P<0.01, by QAP).

**CONCLUSION:** SNA provided a description of patterns of communications in teamwork in an acute medical unit. We used SNA statistics to reveal variation in patterns of team communication and teammate interconnectedness by shift.

## INTRODUCTION

Acute medical unit (AMU) is a collaborative structural organization and system that have emerged to service a demand for immediate and adequate care for life-threatening emergencies. Such high-stake environment needs an efficient communication between the staff members for ensuring patient safety and reducing the risk of adverse effects, as poor communication has been identified as a causal factor in adverse incidents harming the patient 1. Moreover, it has been proved that improvements in communication have decreased medical errors, as well as improved technical performances 2. In medical communication, the quality of information is measured by the verbal and non-verbal content of the actors’ message based on their effective communication skills in the relationship with the patient and the care team. However, the quality of the message spread between actors and the relationships between them is rarely evaluated. Health systems research studies the organization of people, institutions, and resources that deliver health care services to meet the health needs of target populations. A health systems perspective recognizes the inherent complexity of healthcare and the interdependent nature of components within the system. Social network analysis seeks to understand networks and their participants and has two main focuses: the actors and the relationships between them in a specific social context. It defines the social capital in a network, the flow of information, the level of a critical concentration of communication or information and it’s nature, defines specific groups and key elements, and finally dresses the network in a strategic and well-defined structure ^3^,^4^. Social network analysis (SNA) represents a distinctive set of methodologies to map, measure, and analyze social relationships between people, teams, and organizations. It facilitates the exploration of patterns and types of relationship between actors, where these actors (which may be individuals, groups or organizations) are visually represented in a network map by structural nodes, and relationships (ties or links) between these nodes. Using mathematical tools and specialized software packages, researchers can explore how patterns of relationships can operate to facilitate or inhibit communications, actions, and capacities in systems. To our knowledge; this is the first study that traced medical interaction and evaluating the role of each team member on one day in an acute medical unit using social Network Analysis. We sought to demonstrate the value of SNA maps and metrics to understand information sharing. Thus this study aimed to describe the patterns of communications in the teamwork of an acute medical unit (AMU).

## METHODS AND PROCEDURES

Study design: It was a descriptive trans-sectional survey conducted in an Acute Medical Unit (AMU) of Ibn-Sina University Hospital in Rabat, Morocco.

### Study Setting

The AMU unit admits approximately 950 patients annually. Patients are admitted mainly from the emergency unit, falling into a wide range of medical illnesses. Participant: All members of the AMU team who were effective healthcare workers or student affiliated to the AMU at the time of the data collection were included.

### Data collection

Socio-demographic and professional characteristics of health workers included; age, gender (male or female), matrimonial status (married or single). Medical team function included: 3 senior physicians (SP), 3 hospital practitioners (HP), 14 junior physicians (JP), 5 medical students (MS), 16 nurses (RN), a physiotherapist (PHT), 14 healthcare technician HCT), a medical secretary and an archives manager.

### B. SNA INSTRUMENT

We analyzed communication using the Social Network Analysis instrument(SNA).

The social network is defined as a set of social actors or subjects, connected with a set of lines or links representing one or more social relations among them 5. The individual filling out the questionnaire is referred to as “ego”; the contacts that the ego reports on are called “alters”^6^.

A network can be expressed in a form of the graph; displayed in a form of a diagram including nodes connected by links. A network can be either oriented or not oriented. Orientation is the distinction of two separate links between 2 same persons: A to B, and B to A.

A network can be also expressed as a matrix; this creates cells containing information regarding communication between two individuals. The link is read from the actor in the row to the one in the column, giving, therefore, a possibility of orientation with a vector. Therefore, a network is oriented by default. Though reporting a network’s data can either choose to reflect its orientation or not. If in the network the links are not oriented, the matrix is symmetrical. The cell connecting an actor to himself is filled with zero and is not taken into account. In our study, we chose to draw a non-oriented network reporting the frequency of communication considering the maximal digit reported between two individuals making therefore symmetry.

### PROCEDURE

We gathered the list of all AMU staff members and included a total of 58 eligible participants.

Each SNA survey included the names of all eligible AMU personnel. We interviewed every member separately in retrospect of the 24 hours.

Each participant had to report the frequency of communication they established on a scale of 0 being no communication to +10 being more than ten times communication was established. All form of communication was including; face to face interaction; phone call; and a textual message. We followed standard SNA practice and presented a single survey item on its page and included a standard item stem: a) “Think only about the last 24 hours working hours. The record below the total number of times you initiated communication…….when you interact about the health care situation. Health care situation communication included discussion concerning patient care.

We created a sociometric (whole-network) questionnaire. The team’s social network is represented as a square matrix including all team members who completed the questionnaires (rows) and their named discussion partners (columns). Each cell takes the value of the frequency of communication from 0 to +10. We considered the highest frequency reported between two individuals and the reciprocity of the reported interaction, drawing a symmetrical no oriented network. Figure 1 Illustrate the SNA survey and matrices used for calculation of SNA.

**Figure 1:**
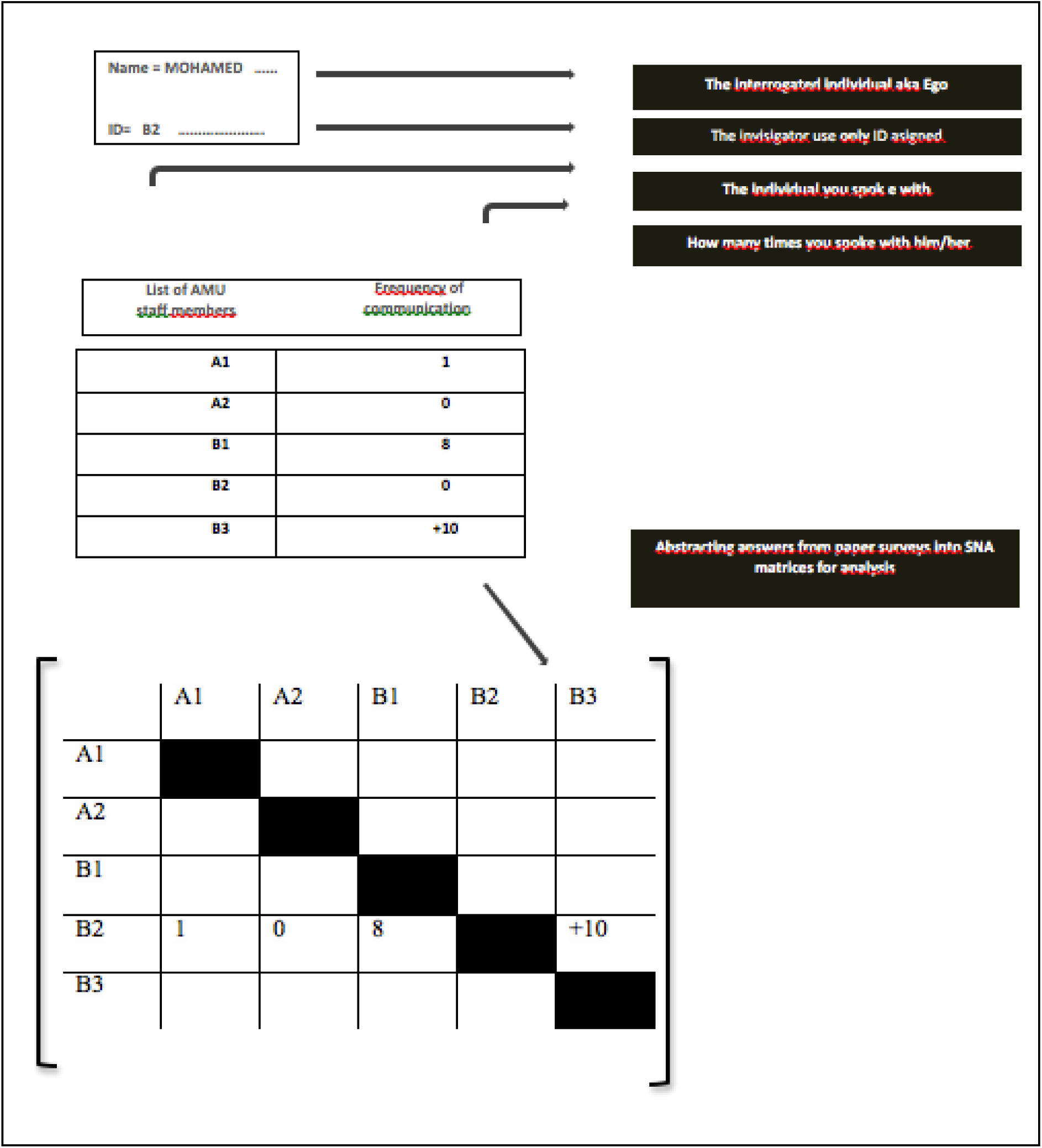
Illustration of SNA survey and matrices used for calculation of SNA Measures in our study

### STATISTICAL METHOD

We used frequencies, percentages, means, and standard deviations to describe the characteristics of participants. Network analysis was used to build network maps using multidimensional scaling and to generate a visual representation of networks using network diagrams. Standard SNA measures included network-level, and nodes level metrics. In this study, nodes represent individuals in the network. ^3^

#### Network-level metrics

provide information about how the network as a whole is operating and included;

**-Network size** represents the number of nodes in the network;

**-*Network density*** is the proportion of existing communication relationships between members (presence or absence) at the dyadic level divided by the total possible number of communication ties at the dyad level. Values for density range from 0 to 100 with higher values indicating greater cohesion and frequent communication among all members

**-*Network Centralization*** The index is valued from 0 to 1. Low centralization (or decentralization) indicates the greater distribution of communication across teammates with no single team member enjoying a high level of communication over any other team member in the network. In contrast, higher values of network centralization indicate that communication is concentrated on one or a selected few members in the team, leaving some members isolated or “out of the loop” ^7^

**-*Network homophily*:** The homophily reflects the degree to which network is composed of subgroups that interact within the groups or with another group. Assuming two groups based on some attribute, one defined as internal and the other as external. We examined the homophily of the network using E-I index = E-I/E+I (from −1to 1);” E” is the number of external friendship links and “I” is the number of internal friendship links. A score of +1 = All links external to subunit, a score of 0 = Links are divided equally, a score of −1 = All links are internal to subunit. P-value was obtained based on 1000 QAP permutations of the quadratic network assignment procedure. The QAP test is robust against this nagging problem of autocorrelation in the network data, removing virtually all the bias that would be introduced with more traditional statistical tests ^8,9^.

#### Node-level metrics ^3^

provide useful information about individuals in the network and can identify who in the network is more or less influential. key node-level metrics used in this study were degree centrality and betweenness centrality.

***-Degree centrality*** ^3^ gives an insight into the rank of the network’s members based on their degree of prominence and influence on the flow of information and network. It ranges from 0 to 1, and a higher value indicated a higher number of direct links to the individual, which provides him with a better positioning level of activity within the network. However, the degree does not capture the node’s location in the larger network, for which we use betweenness centrality.

***-Betweenness centrality*** ^3^: It ranges from 0 to 1 with the higher value indicating a higher number of direct links to the individual. Betweenness centrality refers to the number of times a member connects with other network members that would not otherwise be connected.

The “newcommands” package in Stata 14.2 SE version (Stata Corp, College Station,TX) was used to construct Network maps using multidimensional scaling, and calculate network characteristics ^10^.

## RESULTS

### CHARACTERISTICS OF AMU TEAM MEMBERS

A total of 58 AMU team members were included, 62.1% were women with a mean age of 37±13 years; 46.6% were married. The median distance between hospital and residency was 9 km. Median [IQR] of years of experience was 5.5 [1-26]. Mean ± sd working and sleeping hours were 33.1± 8.7 and 5.6 ± 2.2 respectively.

### SOCIAL NETWORK ANALYSIS

#### Network-Level Metrics

The network size was 58 nodes or actors with 460 connexions. From the resulting map, network metrics indicate a density score of 0.28 (28%.) Figure 2 illustrate AMU network mapping.

**Figure 2:**
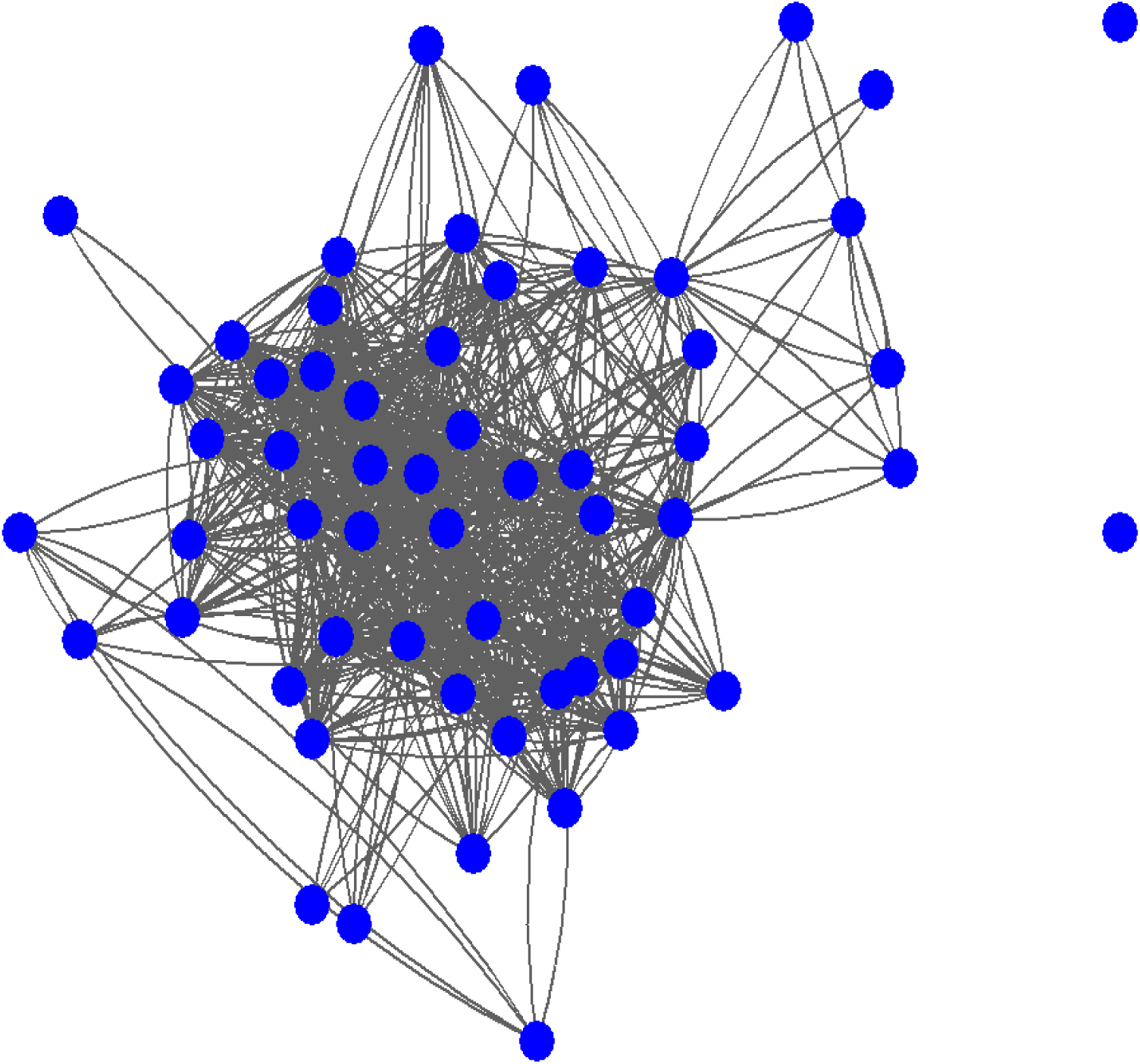
graph of Amu network map

Network centralization was 0.37, indicating a relatively decentralized network.

Network homophily: The E-I index was 0.34 indicating statistically significant heterophily (p=0.01; by QAP).

#### Node-Level Metrics

*Degree centrality*, the network metrics showed that seven team members were central to the network. A centrality degree > 50% included: three senior physicians, the head nurse, the physiotherapist, medical secretary and archives manager. Figures 3 illustrate the AMU node centrality degree;

**Figure 3.**
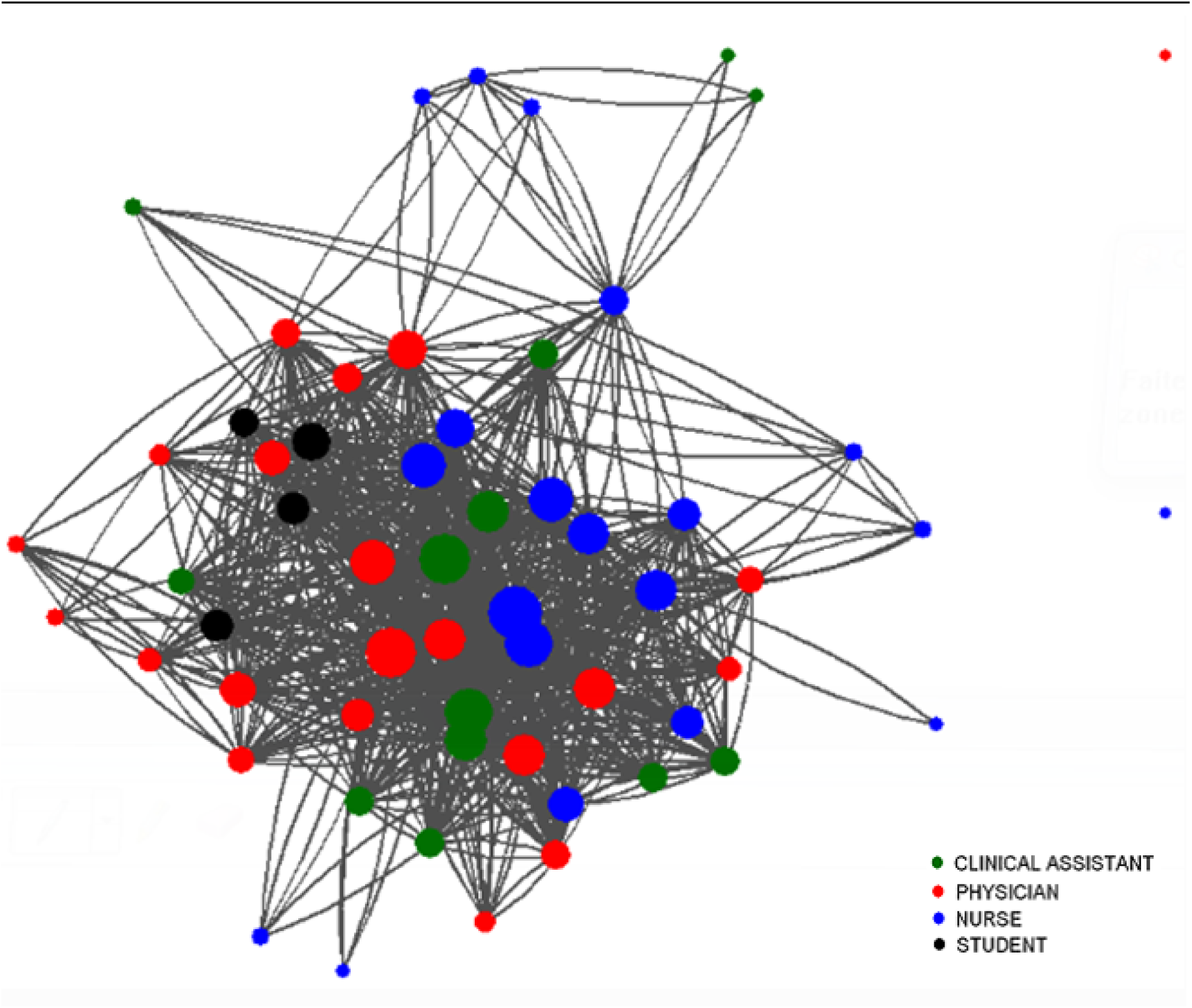
Graph of Amu node-metric: the size of the node is proportional to the percentage of degree centrality; bigger the node indicated higher centrality degree.

Highest *betweenness centrality* was observed in one senior physician, and a junior physician, two nurses and the medical secretary.

## DISCUSSION

This first network analysis in a developing country medical setting highlight the functioning of the team in an acute medical unit and illustrated the patterns of team communication and teammate interconnectedness during 24 last working hours.

The study revealed a moderate team member connectedness as evaluated by density; and a relatively decentralized network, reflecting the stability of the network. Degree centrality identified most active members; represented by three senior physicians, the head nurse, a physiotherapist, medical secretary and archive manager. These members represent the most contacted members. However, one senior physician, one junior physician, two nurses and the medical secretary have a central role in bridging communication as calculated by higher betweenness score.

Our study showed network stability relies on having less chance of collapsing since its communication foundation doesn’t rely on a group of central members. Network metrics showed team member tendency to communicate with the members of different function; suggesting openness and complementarity between the different functions of the team. Nodes metrics showed a homogeneous distribution of interactions between all medical team members; and a central role of all professional profiles within the team suggesting well-functioning organisations. However, no clusters were identified suggesting robust communication patterns.

This study contributes to the literature on health care teams as networks, where little prior work has been published. While limited to brief periods of assessment, Previous research was conducted in the emergency department ^11^ and burn intensive care unit ^[6]^ demonstrated the importance of all team members in the admission of patient and continuity of care.

SNA represents a valuable tool of evaluation and therefore assistance and guidance in the design of an effective quality improvement of teams ^12^. Devoting time and energy in communication and social network analysis is a promising investment in the intent of developing sustainable and well-functioning organizations.

Nonetheless, the retrospective nature of data collection was based on the recall ability of each individual, as well as their honesty with no possible way to verify any answer provided. Though we sought to reduce the recall bias by interviewing the members immediately after the 24 working hours. And finally, the structure of the AMU staff composition and organization may be different from one unit to another, challenging the application on a larger scale and reproduction.

## CONCLUSION

This social network analysis highlights the patterns of team communication and teammate interconnectedness during 24 hours working time in a developing country medical setting. This evaluation revealed network stability since its communication foundation doesn’t rely on a group of central members. Network metrics showed team member tendency to communicate with the members of different function. Nodes metrics showed a homogeneous distribution of interactions between all medical team members; and a central role of all professional profiles within the team suggesting well-functioning organisations. However, no clusters were identified suggesting robust communication patterns.

Such findings can be detrimental to the success of interventions developed. These results increase the awareness of leaders about the power of networks, to further catalyze relationships and connections, and to strengthen the capacity of the network to act collectively. Social network analysis is a promising investment for evaluating and developing sustainable and well-functioning organizations.

## Data Availability

all data referred to the manuscript are available

